# Disentangling neurodegeneration from ageing in multiple sclerosis: the brain-predicted disease duration gap

**DOI:** 10.1101/2024.01.02.23300497

**Authors:** Giuseppe Pontillo, Ferran Prados, Jordan Colman, Baris Kanber, Omar Abdel-Mannan, Sarmad Al-Araji, Barbara Ballenberg, Alessia Bianchi, Alvino Bisecco, Wallace Brownlee, Arturo Brunetti, Alessandro Cagol, Massimiliano Calabrese, Marco Castellaro, Ronja Christensen, Sirio Cocozza, Elisa Colato, Sara Collorone, Rosa Cortese, Nicola De Stefano, Christian Enzinger, Massimo Filippi, Michael A. Foster, Antonio Gallo, Claudio Gasperini, Gabriel Gonzalez-Escamilla, Cristina Granziera, Sergiu Groppa, Yael Hacohen, Hanne F. Harbo, Anna He, Einar A. Høgestøl, Jens Kuhle, Sara Llufriu, Carsten Lukas, Eloy Martinez-Heras, Silvia Messina, Marcello Moccia, Suraya Mohamud, Riccardo Nistri, Gro O. Nygaard, Jacqueline Palace, Maria Petracca, Daniela Pinter, Maria A. Rocca, Alex Rovira, Serena Ruggieri, Jaume Sastre-Garriga, Eva Strijbis, Ahmed Toosy, Tomas Uher, Paola Valsasina, Manuela Vaneckova, Hugo Vrenken, Jed Wingrove, Charmaine Yam, Menno M. Schoonheim, Olga Ciccarelli, James H. Cole, Frederik Barkhof, the MAGNIMS study group

**Affiliations:** Queen Square Multiple Sclerosis Centre, Department of Neuroinflammation, UCL Queen Square Institute of Neurology, University College London, London, United Kingdom; MS Center Amsterdam, Radiology and Nuclear Medicine, Vrije Universiteit Amsterdam, Amsterdam Neuroscience, Amsterdam UMC location VUmc, Amsterdam, The Netherlands; Departments of Advanced Biomedical Sciences and Electrical Engineering and Information Technology, University of Naples “Federico II”, Naples, Italy; Centre for Medical Image Computing, Department of Medical Physics and Biomedical Engineering, University College London, London, United Kingdom; E-Health Center, Universitat Oberta de Catalunya, Barcelona, Spain; Institute of Neuroradiology, St. Josef Hospital, Ruhr-University Bochum, Bochum, Germany; Department of Advanced Medical and Surgical Sciences, University of Campania “Luigi Vanvitelli”, Naples, Italy; Translational Imaging in Neurology (ThINK) Basel, Department of Biomedical Engineering, Faculty of Medicine, University Hospital Basel, University of Basel, Basel, Switzerland; Neurologic Clinic and Policlinic, MS Center and Research Center for Clinical Neuroimmunology and Neuroscience Basel (RC2NB), University Hospital Basel, University of Basel, Basel, Switzerland; Department of Neurosciences, Biomedicine and Movement Sciences, University of Verona, Verona, Italy; Department of Information Engineering, University of Padova, Padova, Italy; Department of Medicine, Surgery and Neuroscience, University of Siena, Siena, Italy; Department of Neurology, Medical University of Graz, Graz, Austria; Neuroimaging Research Unit, Division of Neuroscience, IRCCS San Raffaele Scientific Institute, Neurology Unit, Neurorehabilitation Unit, Neurophysiology Service, IRCCS San Raffaele Scientific Institute, Milan, Italy; Vita-Salute San Raffaele University, Milan, Italy; Department of Neurosciences, San Camillo-Forlanini Hospital, Rome, Italy; Department of Neurology, Focus Program Translational Neuroscience (FTN) and Immunotherapy (FZI), Rhine Main Neuroscience Network (rmn2), University Medical Center of the Johannes Gutenberg University Mainz, Mainz, 55131, Germany; Department of Neurology, Oslo University Hospital, Oslo, Norway; Institute of Clinical Medicine, University of Oslo, Oslo, Norway; Department of Psychology, University of Oslo, Oslo, Norway; Neuroimmunology and Multiple Sclerosis Unit Laboratory of Advanced Imaging in Neuroimmunological Diseases (ImaginEM). Hospital Clinic Barcelona, Fundació de Recerca Clínic Barcelona-Institut d’Investigacions Biomèdiques August Pi i Sunyer and Universitat de Barcelona, Barcelona, Spain; Nuffield Department of Clinical Neurosciences, University of Oxford, Oxford, United Kingdom; Department of Molecular Medicine and Medical Biotechnology, University of Naples “Federico II”, Naples, Italy; Department of Neurosciences and Reproductive and Odontostomatological Sciences, University of Naples “Federico II”, Naples, Italy; Department of Human Neurosciences, Sapienza University of Rome, Rome, Italy; Section of Neuroradiology, Department of Radiology, Hospital Universitari Vall d’Hebron, Universitat Autònoma de Barcelona, Barcelona, Spain; Centre d’Esclerosi Múltiple de Catalunya (Cemcat), Department of Neurology/Neuroimmunology, Hospital Universitari Vall d’Hebron, Universitat Autònoma de Barcelona, Barcelona, Spain; MS Center Amsterdam, Neurology, Vrije Universiteit Amsterdam, Amsterdam Neuroscience, Amsterdam UMC location VUmc, Amsterdam, The Netherlands; Department of Neurology and Center of Clinical Neuroscience, First Faculty of Medicine, Charles University and General University Hospital, Prague, Czech Republic; Department of Radiology, First Faculty of Medicine, Charles University and General University Hospital, Prague, Czech Republic; MS Center Amsterdam, Anatomy and Neurosciences, Vrije Universiteit Amsterdam, Amsterdam Neuroscience, Amsterdam UMC location VUmc, Amsterdam, The Netherlands; Centre for Medical Image Computing, Department of Computer Science, University College London, London, United Kingdom; Dementia Research Centre, UCL Queen Square Institute of Neurology, University College London, London, United Kingdom

**Keywords:** multiple sclerosis, brain ageing, neurodegeneration, disease duration, MRI

## Abstract

Disentangling brain ageing from disease-related neurodegeneration in patients with multiple sclerosis (PwMS) is increasingly topical. The brain-age paradigm offers a window into this problem but may miss disease-specific effects. Here, we statistically modelled disease duration (DD) in PwMS as a function of brain MRI scans and evaluated whether the brain-predicted DD gap (i.e., the difference between predicted and actual duration) could complement the brain-age gap as a DD-adjusted global measure of multiple sclerosis-specific brain damage.

In this retrospective study, we used 3D T1-weighted brain MRI scans of PwMS (i) from a large multicentric dataset (n = 4,392) for age and DD modelling, and (ii) from a monocentric longitudinal cohort of patients with early multiple sclerosis (n = 252 patients, 749 sessions) for clinical validation. We trained and tested a deep learning model based on a 3D DenseNet architecture to predict DD from minimally pre-processed brain MRI scans, while age predictions were obtained with the previously validated DeepBrainNet algorithm. Model predictions were scrutinised to assess the influence of lesions and brain volumes, while the DD gap metric was biologically and clinically validated within a linear model framework assessing its relationship with brain-age gap values and with physical disability measured with the Expanded Disability Status Scale (EDSS).

Our model predicted DD better than chance (mean absolute error = 5.63 years, R^2^ = 0.34) and was nearly orthogonal to the brain-age model, as suggested by the very weak correlation between DD gap and brain-age gap values (*r* = 0.06). DD predictions were influenced by spatially distributed variations in brain volume, and, unlike brain-predicted age, were sensitive to the presence of lesions (mean difference between unfilled and filled scans: 0.55 ± 0.57 years, *p* < 0.001). The DD gap metric significantly explained EDSS scores (β = 0.060, *p* < 0.001), adding to brain-age gap values (ΔR^2^ = 0.012, *p* < 0.001). Longitudinally, increasing annualised DD gap was associated with greater annualised EDSS changes (*r* = 0.50, *p* < 0.001), with a significant incremental contribution in explaining disability worsening compared to changes of the brain-age gap alone (ΔR^2^ = 0.064, *p* < 0.001).

The brain-predicted DD gap metric appears to be sensitive to multiple sclerosis-related lesions and brain atrophy, adding to the brain-age paradigm in explaining physical disability both cross-sectionally and longitudinally. Potentially, it may be used as a multiple sclerosis-specific biomarker of disease severity and progression.

## Introduction

In multiple sclerosis, a complex interplay exists between brain ageing and disease-related tissue damage accumulation.^1^ Untangling the shared and unique aspects of ageing and multiple sclerosis-related neurodegeneration is important to accurately assess disease severity and progression over time, and is increasingly relevant as both life expectancy and the average age of patients with multiple sclerosis (PwMS) are increasing.^2^ However, measuring the two processes independently is an open challenge due to their substantial overlap and dynamic interaction.^3^

The brain-age paradigm has emerged as a promising data reduction strategy, summarizing complex neuroimaging information into a simple yet clinically relevant biomarker of ageing and neurodegeneration.^4^ Briefly, machine learning methods are used to model chronological age as a function of brain MRI scans in healthy subjects (HS), and the resulting model of normal brain ageing is used for neuroimaging-based age prediction in unseen subjects.^4^ The extent to which a subject deviates from healthy brain ageing, expressed as the difference between predicted and chronological age (the brain-age gap, BAG), has been proposed as an age-adjusted global index of brain health, capturing variations associated with a wide spectrum of neurological and psychiatric disorders, including multiple sclerosis.^5–7^

However, the BAG metric is designed to be sensitive to those aspects of brain pathology that most resemble healthy ageing processes, potentially failing to capture disease-specific effects. Indeed, conceptualizing brain involvement in multiple sclerosis solely as a form of premature/accelerated ageing might be reductive since it differs from healthy brain ageing not only in grade but also in nature. Brain volume loss, for instance, is known to occur with different spatio-temporal patterns in healthy ageing and multiple sclerosis^8^, while white matter (WM) lesions are characteristic of multiple sclerosis but do not substantially determine brain-age prediction^7^. Furthermore, BAG is influenced by early-life genetic and environmental factors which may not be intrinsically related to ageing processes, nor to the development of brain pathology.^9^

While there is increasing attention to the prodromal and pre-clinical aspects of multiple sclerosis, a discrete clinical onset date is almost always identifiable in PwMS which, although inherently ambiguous, might represent an acceptable proxy for disease start and enable the estimation of disease duration (DD).^10^ Interestingly, this has been previously used to contextualize individual disease severity in PwMS by referencing a clinical indicator (e.g., the Expanded Disability Status Scale - EDSS - score) to its distribution in patients with comparable DD (i.e., the Multiple Sclerosis Severity Score - MSSS).^11^

Here, we proposed a quantitative neuroimaging measure of brain structural damage, assessed through conventional MRI and referenced to DD. We hypothesised that modelling DD in PwMS as a function of structural brain MRI scans would provide a reference standard of multiple sclerosis-related brain damage accumulation. The error associated with the prediction of DD (the brain-predicted DD gap), quantifying the extent to which a patient deviates from the typical disease trajectory, should reflect past and ongoing multiple sclerosis-specific processes and encode biologically and clinically relevant information about disease-related variability. By evaluating it against BAG and longitudinal clinical and MRI data, we aimed to validate the brain-predicted DD gap as a neuroimaging biomarker of multiple sclerosis severity and progression.

Finally, as multiple sclerosis is sometimes theorized as a purely age-dependent disease, with natural history driven by age irrespective of the apparent DD,^12^ we also explored an alternative modelling strategy originating from this alternative conceptualization of the disease. Specifically, we modelled chronological age in PwMS to estimate a reference trajectory of multiple sclerosis-specific brain ageing (MS-age) and tested the corresponding prediction error (the brain-predicted MS-age gap) as a biomarker of disease severity and progression.

## Materials and methods

### Participants

In this retrospective multi-centric study, we collected MRI and clinico-demographic data of patients diagnosed with multiple sclerosis according to the 2010 McDonald criteria^13^ or clinically isolated syndrome (CIS)^10^. Written informed consent was obtained from each participant independently at each centre. The final protocol for this study was reviewed and approved by the local Ethics Committees and the MAGNIMS Study Group Steering Committee for the analysis of pseudonymized data (www.magnims.eu).

We gathered 3D T1-weighted (T1w) brain MRI scans of patients from 15 European centres for modelling age and DD. A T2-weighted fluid-attenuated inversion recovery (FLAIR) scan was also required for all subjects to automatically segment T2-hyperintense lesions. For the clinical and biological validation of the age and DD models, we used 3D T1w brain images of an external longitudinal cohort of patients with a first scan in the early phases of the disease (< 5 years from clinical onset). Physical disability was scored using EDSS at the time of MRI.

### Deep learning age and disease duration modelling

A schematic illustration of the conceptual design of the study is shown in Figure 1.

**Figure 1.**
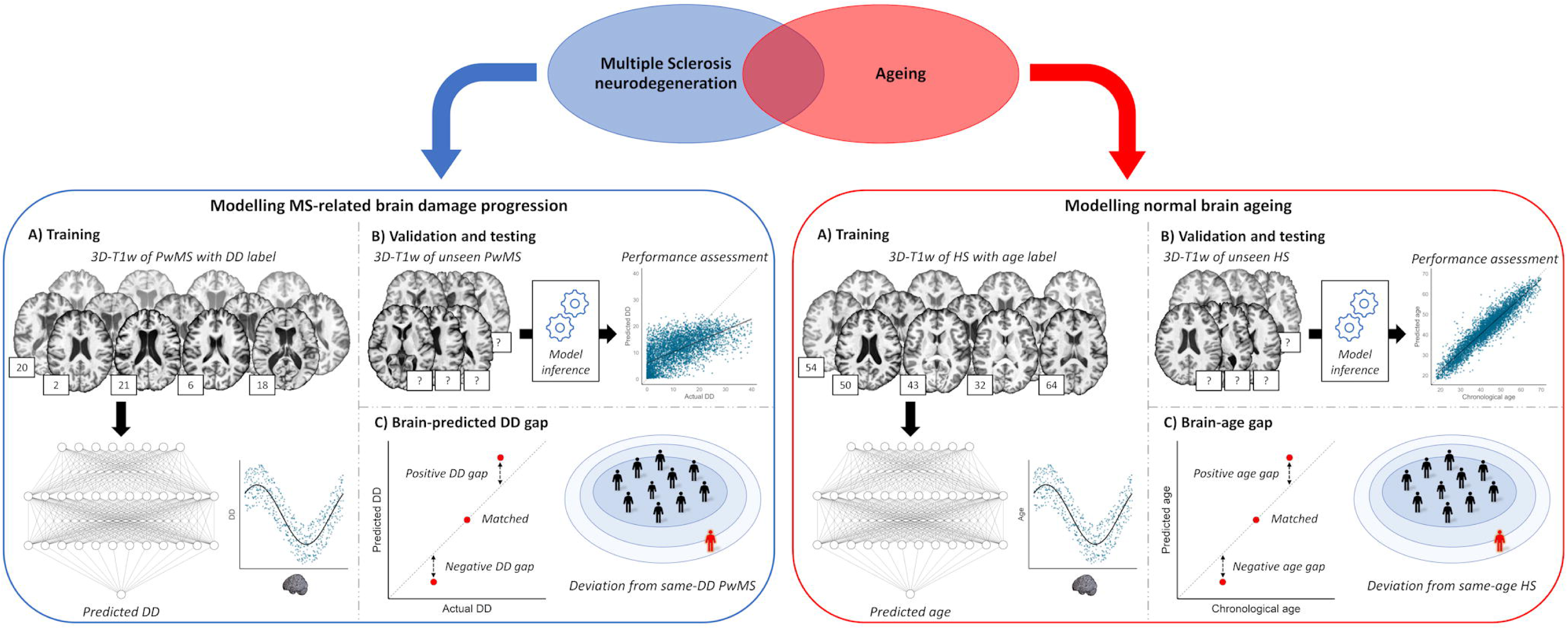
Conceptual framework of the study. With the brain-age paradigm, chronological age is modelled as a function of brain MRI scans in healthy subjects, and the resulting model of normal brain aging is used for neuroimaging-based age prediction in unseen subjects. The extent to which a subject deviates from healthy brain ageing, expressed as the difference between predicted and chronological age (the brain-age gap), is an age-adjusted global index of brain health. We proposed to complement this approach by additionally modelling DD in PwMS as a function of brain MRI scans, to provide a reference standard of multiple sclerosis-related brain damage accumulation. The error associated with the prediction of DD (the brain-predicted DD gap), quantifying the extent to which a patient deviates from the typical disease trajectory, is a DD-adjusted global measure of multiple sclerosis-specific brain damage.

T1w scans were used as input for age and DD prediction models. Minimal preprocessing was performed with ANTsPyNet (https://github.com/ANTsX/ANTsPyNet) and included N4 bias field correction, skull-stripping and affine registration to the 1mm-isotropic MNI152 template.

For the prediction of age and the estimation of BAG, we used the DeepBrainNet algorithm, an external model based on a two-dimensional convolutional neural network architecture that has been extensively validated.^14^ The BAG model was considered as a benchmark in subsequent analyses.

For the prediction of DD, an in-house model was trained, validated, and tested on the age and DD modelling cohort, which was randomly split into training (n = 2811, 64%), validation (n = 703, 16%) and test (n = 878, 20%) sets. Our model was built on the 3D DenseNet264 architecture, adapted from the implementation available at Project MONAI (https://docs.monai.io/en/stable/_modules/monai/networks/nets/densenet.html) by adding a linear regression layer for the prediction of a continuous variable and a 0.2 dropout rate after each dense layer to reduce overfitting. Before being presented to the model, images were resampled to 1.5 mm^3^ voxels to reduce array size and computational burden while retaining anatomical details, and online data augmentation was performed, including random spatial and intensity transformations, to make the network further invariant to image quality variations and site effects. A log(x+1) transformation was applied to the outcome variable to account for the highly positively skewed distribution of DD values. Mean absolute error (MAE) and coefficient of determination (R^2^) were used to quantify model performance. Modelling was performed with PyTorch 1.12.0^15^ using one NVIDIA Tesla T4 16 GB graphics processing unit.

Finally, to model multiple sclerosis-specific brain-ageing, we trained, validated and tested the same network architecture to predict chronological age on the age and DD modelling cohort. A conceptual outline of the MS-age modelling strategy is shown in Supplementary Figure 1.

### Model interpretability

To scrutinise model predictions on the test set, we used guided backpropagation to obtain saliency maps highlighting regions of the input image that are most influential for the model’s predictions.^16^ Furthermore, to better understand the imaging patterns underlying the predictions, we conducted a post-hoc correlational analysis: we segmented T1w and FLAIR scans of the test set with SAMSEG^17^ and FastSurfer^18^ to obtain volumes of lesions and gray matter regions, respectively, which were then correlated with age and DD gaps while correcting for age, age^2^, DD, sex, and estimated total intracranial volume (eTIV).

Finally, we investigated the impact of MS lesions on age and DD predictions: lesions were artificially removed from T1w images of the test set using FSL lesion-filling algorithm^19^, both “lesion-filled” and “unfilled” scans were run through the prediction procedures, and resulting values were compared with paired sample t-tests and Bland-Altman plots.

### Statistical analysis

Statistical analyses were carried out using R (version 4.1.2), with a statistical significance level set at *p* < 0.05. Cross-sectional associations between age and DD gaps and EDSS were investigated in the test set using linear models including also age, age^2^ (to account for the non-linear effect of age), DD, and sex. To assess the additional values of the DD and MS-age gap metrics over the “classical” BAG in explaining EDSS variance, the corresponding models were compared using F tests.

In the early multiple sclerosis cohort, the longitudinal evolutions of EDSS, BAG, DD and MS-age gaps were analysed using a multilevel linear model framework, with timepoints nested within subjects and random intercept and slope of follow-up time per subject, including also the fixed effects of age, age^2^, and sex. When modelling the DD gap, the fixed effect of DD was also included in the model to correct for DD-related bias (i.e., the underestimation of DD in long-standing PwMS and vice versa). From these growth models, individualized changes per year (i.e., annualised) were extracted as the individual-level coefficients of the follow-up time term, corresponding to the sum of the fixed and random effects. Then, we explored how longitudinal changes in brain MRI-derived measures related to changes in physical disability (i.e., EDSS) by correlating the corresponding annualised changes in patients with at least two visits (n = 200). Similar to the cross-sectional analysis, the value of adding the longitudinal evolution of DD or MS-age gap to BAG change over time for explaining EDSS worsening was assessed by comparing the corresponding models with F tests.

## Data availability

Data from patients are controlled by the respective centres (listed in Table 1) and are therefore not publicly available. Request to access the data should be forwarded to data controllers via the corresponding author. The trained DD and MS-age models will be made available at https://github.com/giupontillo.

**Table 1.** Demographic and clinical characteristics of the studied population.

| Cohort | N | Age |  |  | DD |  |  | N | EDSS |  | CIS / RR / SP / PP / NA |
| --- | --- | --- | --- | --- | --- | --- | --- | --- | --- | --- | --- |
|  |  | Mean | SD | Range | Mean | SD | Range |  | Median | Range |  |
| **Age and DD modelling** | 4,392 | 42.8 | 10.6 | 16.4 - 72.8 | 11.4 | 9.3 | 0.0 - 51.6 | 4,341 | 2.0 | 0.0 - 7.5 | 134 / 2945 / 347 / 81 / 885 |
| Barcelona I | 885 | 44.3 | 9.9 | 18.4 - 70.6 | 13.5 | 8.7 | 0.0 - 45.8 | 878 | 2.0 | 0.0 - 6.5 | 0 / 0 / 0 / 0 / 885 |
| Barcelona II | 61 | 48.9 | 10.4 | 25.6 - 72.2 | 19.7 | 9.5 | 8.4 - 46.1 | 61 | 2.5 | 1.0 - 6.5 | 0 / 52 / 9 / 0 / 0 |
| Basel | 100 | 46.3 | 13.6 | 18.2 - 71.9 | 14.6 | 12.0 | 0.5 - 47.9 | 100 | 3.0 | 0.0 - 7.0 | 1 / 73 / 18 / 8 / 0 |
| Bochum | 101 | 34.2 | 10.7 | 18.4 - 59.5 | 0.7 | 0.7 | 0.0 - 2.5 | 101 | 1.5 | 0.0 - 5.0 | 52 / 49 / 0 / 0 / 0 |
| Graz | 143 | 40.3 | 10.0 | 20.1 - 69.0 | 10.6 | 8.3 | 0.4 - 39.1 | 143 | 1.0 | 0.0 - 7.0 | 3 / 127 / 10 / 3 / 0 |
| Mainz | 350 | 34.8 | 10.5 | 17.0 - 69.0 | 2.8 | 4.6 | 0.0 - 33.0 | 347 | 1.0 | 0.0 - 6.5 | 64 / 285 / 0 / 1 / 0 |
| Milan | 64 | 43.6 | 10.5 | 22.5 - 62.9 | 11.7 | 10.3 | 0.0 - 39.0 | 64 | 4.5 | 1.0 - 7.5 | 0 / 30 / 27 / 7 / 0 |
| Naples I | 220 | 40.0 | 12.6 | 16.4 - 72.3 | 10.4 | 8.5 | 0.0 - 39.4 | 220 | 3.5 | 0.0 - 7.5 | 2 / 158 / 36 / 24 / 0 |
| Naples II | 63 | 37.9 | 10.8 | 20.9 - 61.7 | 9.0 | 8.4 | 0.1 - 32.3 | 63 | 2.0 | 0.0 - 6.5 | 1 / 52 / 5 / 5 / 0 |
| Oslo | 401 | 38.5 | 10.2 | 18.5 - 68.3 | 4.7 | 6.2 | 0.0 - 36.9 | 372 | 2.0 | 0.0 - 7.0 | 7 / 377 / 11 / 6 / 0 |
| Oxford | 16 | 44.8 | 6.5 | 31.9 - 56.0 | 10.8 | 4.7 | 1.8 - 19.5 | 16 | 2.0 | 0.0 - 6.0 | 0 / 16 / 0 / 0 / 0 |
| Prague | 1,785 | 45.4 | 8.9 | 20.1 - 72.8 | 14.0 | 8.9 | 0.0 - 51.6 | 1,774 | 2.5 | 0.0 - 7.5 | 0 / 1540 / 226 / 19 / 0 |
| Rome | 105 | 43.1 | 11.3 | 17.7 - 65.3 | 10.7 | 8.7 | 0.7 - 36.9 | 105 | 2.0 | 0.0 - 6.5 | 1 / 99 / 3 / 2 / 0 |
| Siena | 47 | 44.0 | 12.0 | 17.9 - 71.3 | 12.7 | 8.4 | 1.0 - 38.4 | 47 | 1.5 | 0.0 - 6.5 | 0 / 42 / 0 / 5 / 0 |
| Verona | 51 | 40.0 | 11.4 | 20.3 - 65.5 | 9.1 | 8.7 | 0.0 - 34.9 | 50 | 2.0 | 0.0 - 7.0 | 3 / 45 / 2 / 1 / 0 |
| **Early multiple sclerosis** | 252 | 34.5 | 8.3 | 19.4 - 64.5 | 0.7 | 1.2 | 0.0 - 4.5 | 231 | 1.0 | 0.0 - 6.5 | 110 / 142 / 0 / 0 / 0 |
| London I | 160 | 35.2 | 8.8 | 19.4 - 64.5 | 1.0 | 1.5 | 0.0 - 4.5 | 150 | 1.0 | 0.0 - 6.5 | 49 / 111 / 0 / 0 / 0 |
| London II | 92 | 33.1 | 7.2 | 19.9 - 53.7 | 0.2 | 0.3 | 0.0 - 3.1 | 81 | 1.0 | 0.0 - 3.5 | 61 / 31 / 0 / 0 / 0 |
|  |  | Mean | SD | Range | Mean | SD | Range |  | Median | Range |  |

*Note.* SD = standard deviation; DD = disease duration; CIS = clinically isolated syndrome; RR = relapsing-remitting; SP = secondary-progressive; PP = primary-progressive; NA = not available.

## Results

### Participants

A total of 4,392 patients were selected for the Age and DD modelling cohort. The Early multiple sclerosis longitudinal validation cohort was composed of 252 patients and 749 sessions, with a mean follow-up time of 4.5 years (range: 0.2 - 19.1). Demographic and clinical characteristics of the studied population are reported in Table 1, while the details of the different acquisition protocols are provided in Supplementary Table 1.

### Age and disease duration models

The DeepBrainNet model predictions on the test set showed that multiple sclerosis was associated with older appearing brains (mean BAG: 7.81 ± 9.35 years). As for the prediction of DD, the out-of-sample performance of the model was well above chance level (test set MAE = 5.63 years, R^2^ = 0.34) (Figure 2A). To help contextualize the performance of the DD prediction model, we computed R^2^ values for the association of DD with established MRI biomarkers in the test set for comparison: total lesion (R^2^ = 0.10) and thalamic (R^2^ = 0.14) volumes. As for the MS-age model, predictions on the test set were highly accurate (MAE = 3.78 years, R^2^ = 0.80) (Supplementary Figure 2A).

**Figure 2.**
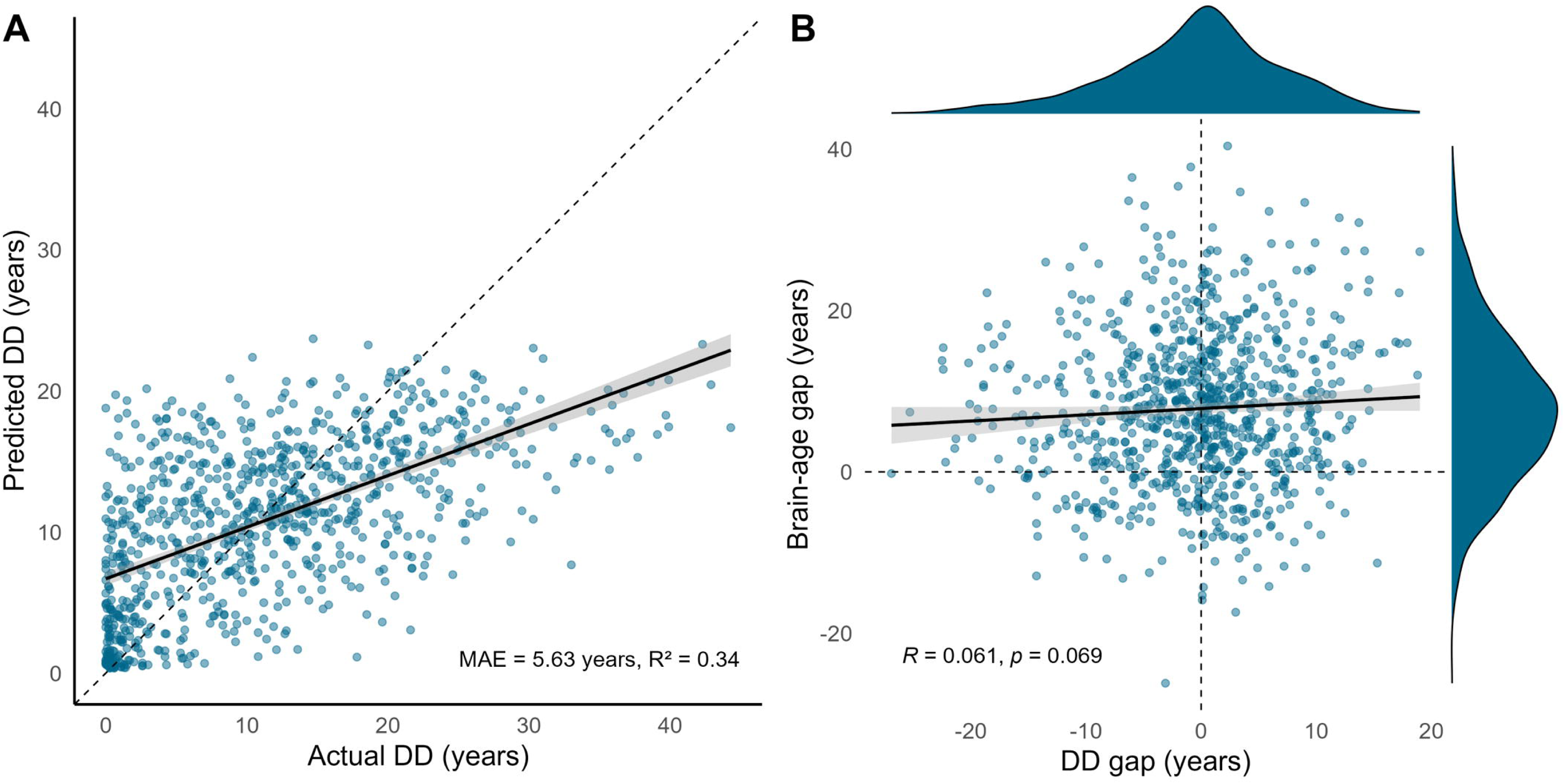
Modelling disease duration in patients with multiple sclerosis. In (A), scatterplot showing the relationship between the actual disease duration values in the test set (N = 878) and the ones predicted by the model. In (B), scatterplot showing the relationship between the disease duration gap and the brain-age gap (obtained with the DeepBrainNet model) in the test set; marginal density plots are also shown, portraying the distribution of the two variables. Linear fit lines are shown as solid lines (with corresponding 95% confidence intervals in grey), while dashed lines represent the line of identity (A), and horizontal and vertical zero reference lines (B), respectively.

When looking at the relationship between these metrics, DD gap and BAG values were nearly orthogonal to each other (*r* = 0.06, *p* = 0.07), suggesting that the models’ predictions were largely independent (Figure 2B). On the other hand, MS-age gap still moderately correlated with BAG (*r* = 0.35, *p* < 0.001), revealing a higher degree of entanglement between the two models (Supplementary Figure 2B).

The interpretability analysis showed that both the DD and MS-age models focused on regions that appear to be primarily related to (the widening of) the cerebrospinal fluid spaces (Figure 3 and Supplementary Figure 3). Also, all age and DD gap measures correlated diffusely with regional brain atrophy and lesion burden, with the greatest effect sizes observed for BAG values (and the lowest for the MS-age gap) and no clear anatomical specificity (Figure 4 and Supplementary Figure 4). As for the impact of MS lesions, there was a significant impact of the filling procedure on brain-predicted DD values (mean difference between unfilled and filled scans: 0.55 ± 0.57 years, *p* < 0.001) (Figure 5A), with no evident systematic bias caused by lesion filling for brain-age predictions (mean difference: −0.03 ± 0.80 years, p = 0.31) (Figure 5B). The MS-age model was also slightly sensitive to the filling procedure (mean difference: 0.07 ± 0.43 years, *p* < 0.001) (Supplementary Figure 5).

**Figure 3.**
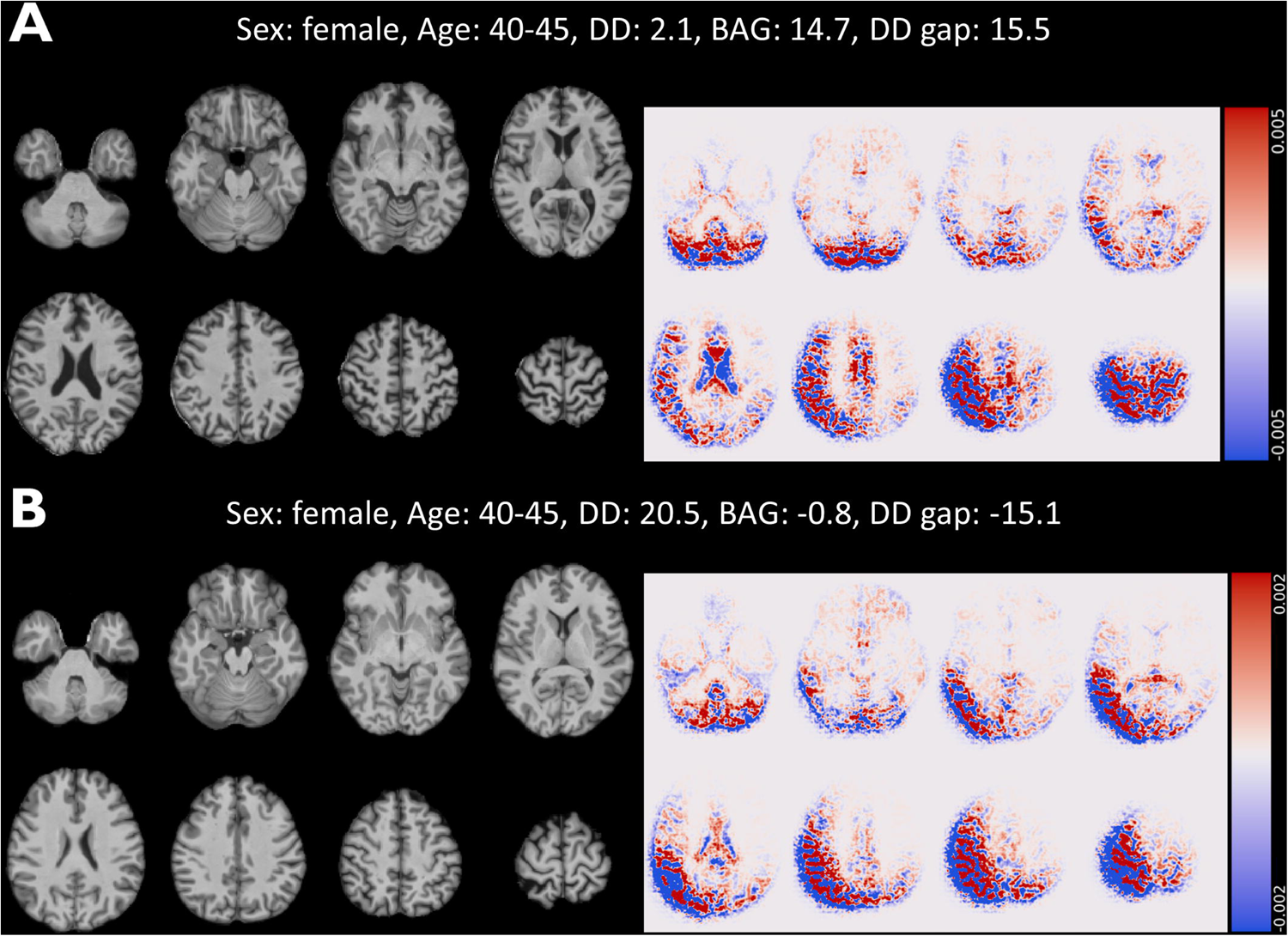
Guided backpropagation analysis to interrogate brain regions influencing the model for the prediction of disease duration. Lightbox view of selected slices from the quasi-raw T1w volumes (on the left) and corresponding guided backpropagation-derived saliency maps (on the right) of two representative pwMS exhibiting extremely positive (A) or negative (B) values of DD gap. For saliency maps, both positive (positively correlated with the output, in *red*) and negative (negatively correlated with the outcome, in *blue*) magnitudes are shown. In both cases, the model focuses mostly on regions that appear to be related to (the widening of) the cerebrospinal fluid spaces.

**Figure 4.**
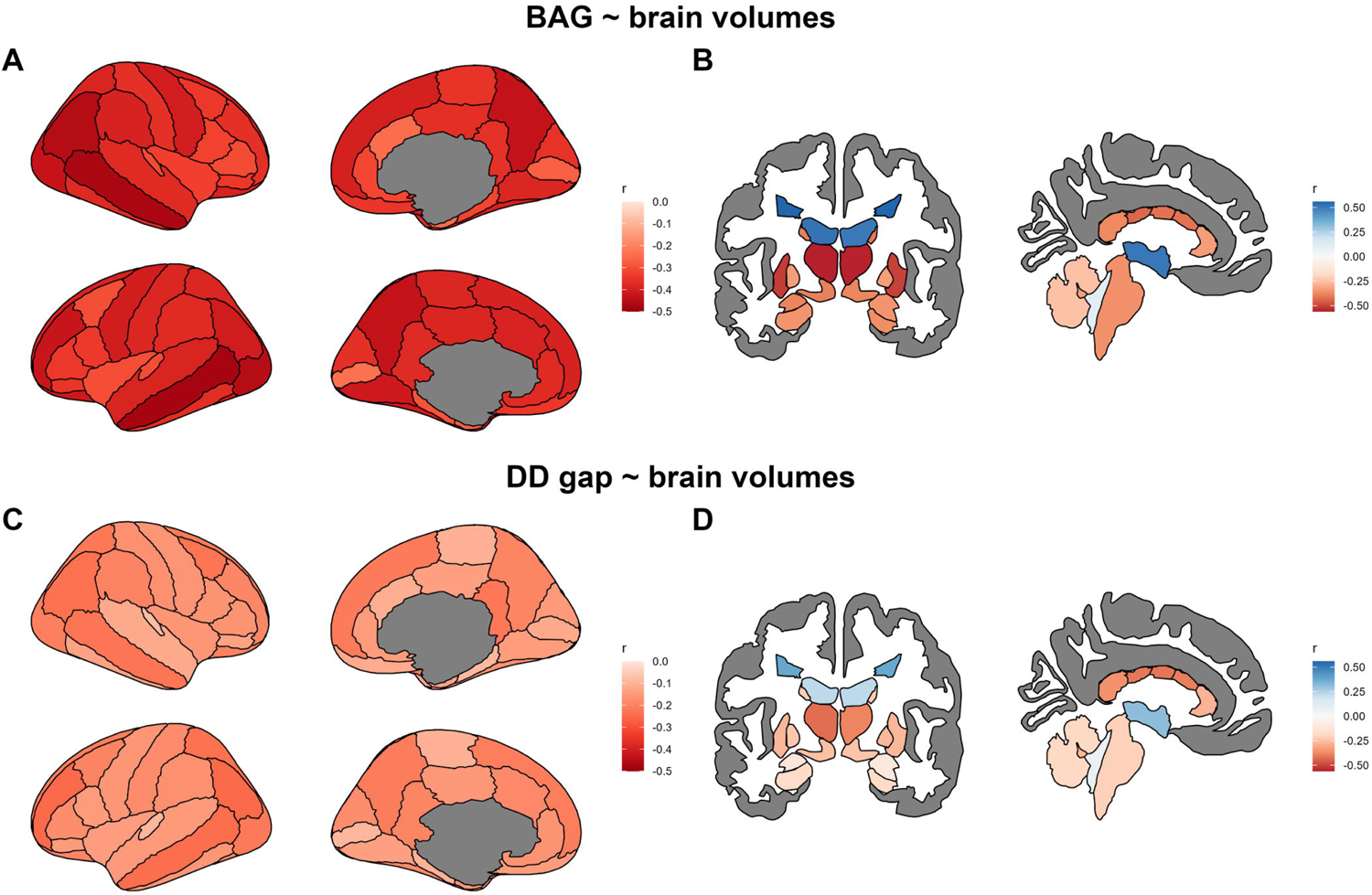
Correlations between brain-age and disease duration gaps and regional brain and lesion volumes. In the upper row, plots showing the correlations between brain-age gap values and cortical (A) and subcortical/lesion (B) volumes. In the bottom row, plots showing the correlations between disease duration gap values and cortical (C) and subcortical/lesion (D) volumes. Shown are the Pearson correlation coefficients resulting from partial correlation analyses correcting for age, age^2^, disease duration, sex, and estimated total intracranial volume. The cortex is parcellated according to the DKT atlas.^18^

**Figure 5.**
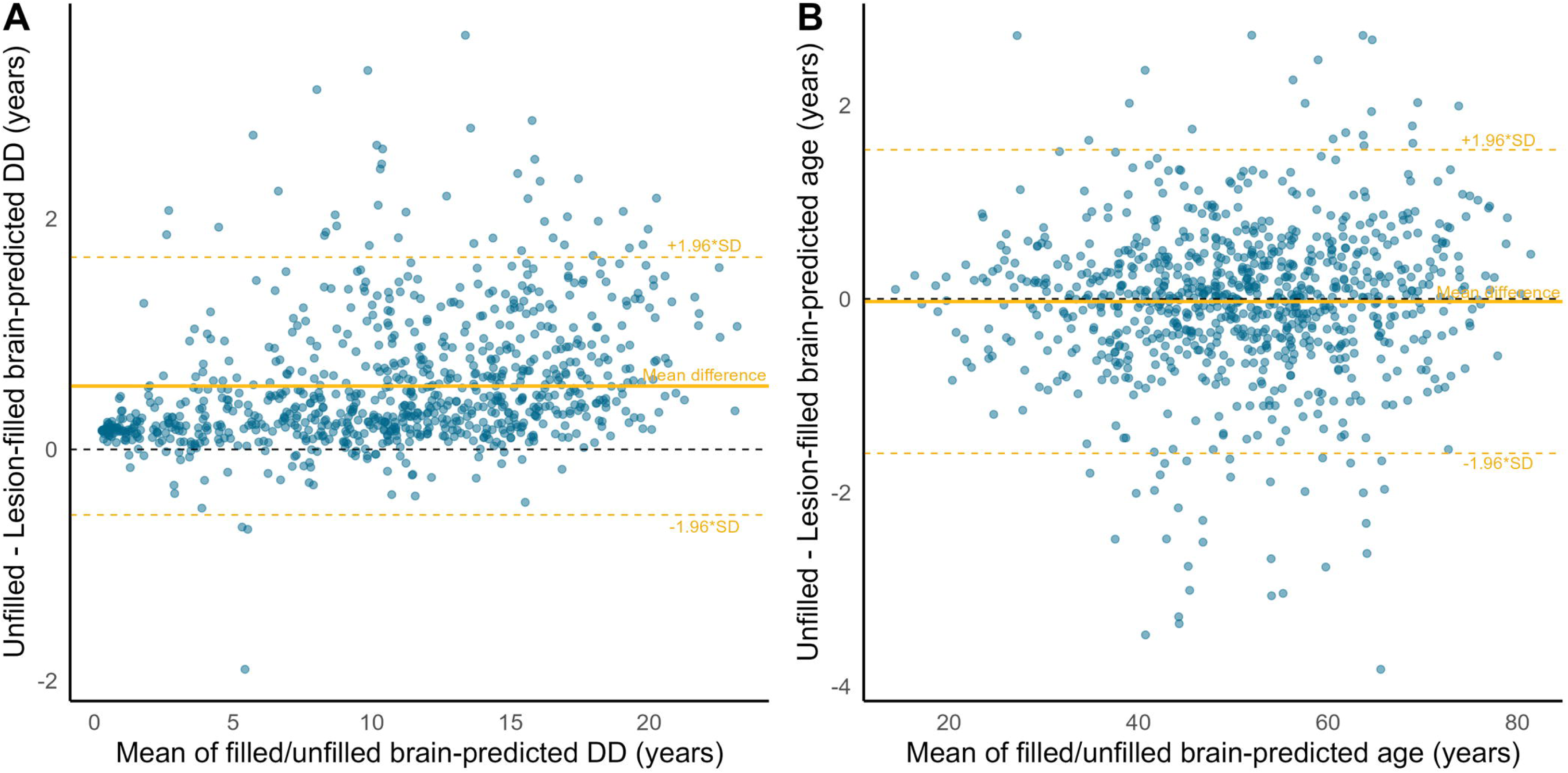
Impact of MS lesions on age and DD predictions. Bland–Altman plot of brain-predicted DD (A) and age (B) from unfilled and filled T1w scans. The plots show the mean value from the 2 measures for each participant (x-axis) and the difference between the 2 measures (y-axis). The mean difference lines are solid, and the corresponding limits of agreement (±1.96 * standard deviation of difference) are dashed lines.

### Brain-age and disease duration gaps independently explain physical disability

In the test set, both BAG (β = 0.026, *p* < 0.001) and DD gap (β = 0.060, *p* < 0.001) were positively associated with EDSS (Table 2 and Figure 6A-B). A positive association was also found for the MS-age gap metric (β = 0.031, *p* < 0.001) (Supplementary Table 2 and Supplementary Figure 6).

**Figure 6.**
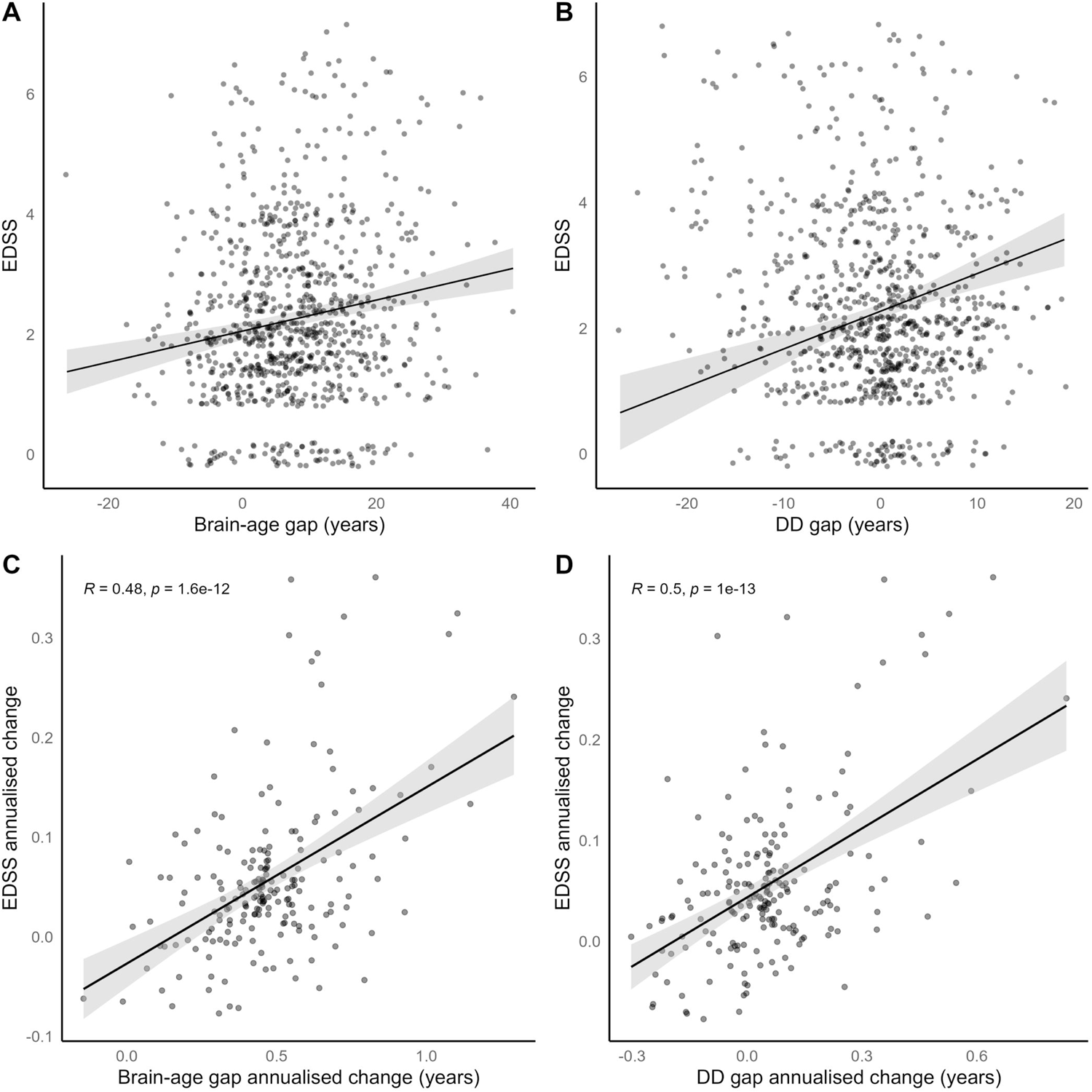
Relationships between brain-age and disease duration gaps and physical disability. In the upper row, scatterplots showing the marginal effects on EDSS of the brain-age (A) and disease duration (B) gap metrics (regression models were corrected for the effects of age, age^2^, disease duration, and sex). In the bottom row, scatterplots showing the relationship between annualised changes of EDSS and brain-age (C) and disease duration (D) gaps. Linear fit lines are shown as solid lines (with corresponding 95% confidence intervals in grey).

**Table 2.** Models for the prediction of EDSS and EDSS annualized change. Coefficient estimates (with standard errors in parentheses), and model statistics for the linear regression analyses predicting EDSS and EDSS annualized change.

|  | *Dependent variable:* |  |  |
| --- | --- | --- | --- |
|  | **EDSS** |  |  |
|  | **(1)** | **(2)** | **(3)** |
| Brain-age gap | 0.026***<br>(0.005) |  | 0.017***<br>(0.006) |
| DD gap |  | 0.060***<br>(0.011) | 0.044***<br>(0.012) |
| Age | 13.786***<br>(1.656) | 6.927***<br>(1.754) | 9.756***<br>(1.988) |
| Age<sup>2</sup> | 2.093<br>(1.337) | 3.234**<br>(1.356) | 3.022**<br>(1.352) |
| DD | 0.023***<br>(0.006) | 0.078***<br>(0.010) | 0.060***<br>(0.012) |
| Sex | -0.094<br>(0.096) | -0.001<br>(0.095) | -0.053<br>(0.096) |
| Constant | 1.900***<br>(0.091) | 1.475***<br>(0.129) | 1.558***<br>(0.131) |
| Observations | 867 | 867 | 867 |
| R<sup>2</sup> | 0.187 | 0.191 | 0.199 |
| Adjusted R<sup>2</sup> | 0.182 | 0.186 | 0.193 |
| Residual Std. Error | 1.318 (df = 861) | 1.315 (df = 861) | 1.309 (df = 860) |
| F Statistic | 39.531*** (df = 5; 861) | 40.538*** (df = 5; 861) | 35.568*** (df = 6; 860) |
|  | **EDSS annualised change** |  |  |
|  | **(1)** | **(2)** | **(3)** |
| Brain-age gap annualised change | 0.177***<br>(0.023) |  | 0.099***<br>(0.029) |
| DD gap annualised change |  | 0.228***<br>(0.028) | 0.150***<br>(0.036) |
| Constant | -0.027**<br>(0.012) | 0.044***<br>(0.005) | 0.001<br>(0.013) |
| Observations | 195 | 195 | 195 |
| R<sup>2</sup> | 0.228 | 0.250 | 0.292 |
| Adjusted R<sup>2</sup> | 0.224 | 0.246 | 0.285 |
| Residual Std. Error | 0.069 (df = 193) | 0.068 (df = 193) | 0.067 (df = 192) |
| F Statistic | 57.158*** (df = 1; 193) | 64.325*** (df = 1; 193) | 39.583*** (df = 2; 192) |

Note: DD = disease duration; df = degrees of freedom. \* $p < 0.1$ ; \*\* $p < 0.05$ ; \*\*\* $p < 0.01$

When investigating the incremental value of multiple sclerosis-specific metrics in explaining EDSS in addition to BAG, the DD gap significantly added to the baseline model (ΔR^2^ = 0.012, *p* < 0.001) (Table 2), while the inclusion of the MS-age gap did not significantly improve the model fit (ΔR^2^ = 0.002, p = 0.10) (Supplementary Table 2).

### Longitudinal brain-age and disease duration gaps’ increases independently explain EDSS worsening

In the early multiple sclerosis cohort, growth models revealed significant EDSS worsening (β = 0.058, *p* < 0.001) and BAG increase (β = 0.472, *p* < 0.001) over time (Supplementary Figure 6). Both DD (β = 0.057, *p* = 0.22) and MS-age (β = 0.016, *p* = 0.76) gaps only exhibited a slight, non-significant, upward trend (Supplementary Figure 7).

The annualised change in EDSS score correlated with annualised changes of both BAG (*r* = 0.48, *p* < 0.001) and DD gap (*r* = 0.50, *p* < 0.001) (Figure 6C-D), while the correlation between annualised changes of EDSS and MS-age gap did not reach statistical significance (*r* = 0.11, *p* = 0.12) (Supplementary Figure 8).

When assessing how longitudinal changes of multiple sclerosis-specific metrics contributed to explaining EDSS worsening, the DD gap significantly added to the baseline model including BAG change over time (ΔR^2^ = 0.064, *p* < 0.001) (Table 2), while the addition of the MS-age gap metric did not significantly improve the model fit (ΔR^2^ = 0.010, p = 0.13) (Supplementary Table 4).

## Discussion

Using deep learning, we separately modelled ageing and disease-specific effects from structural brain MRI scans of a large multicentric sample of PwMS. We validated the DD gap as a biologically and clinically meaningful global measure of multiple sclerosis-specific brain damage, adding to the information provided by models of healthy brain ageing as a biomarker of disease severity and progression.

While age is often treated as a mere confounder in neuroimaging analyses, brain ageing and multiple sclerosis are intimately intertwined. On the one hand, the relationship between age and the brain is shaped by the disease and encodes disease-related information. On the other, age is an essential modifier of multiple sclerosis clinical course and treatment response.^20,21^ Understanding the complex interaction between ageing and neurodegeneration and disentangling the overlapping and distinct mechanisms underlying the two processes bears great transdiagnostic relevance and is the topic of increasing research interest.^22,23^

The brain-age paradigm offers a window into this problem and has been previously used to characterize neurodegeneration in multiple sclerosis due to its sensitivity to brain ageing-like patterns.^5–7,24,25^ In line with previous studies, our results confirmed that, when observed through the lens of healthy ageing, the brains of PwMS look older than normal (around eight years on average), suggesting that at least some of the disease-related variance in brain structure can be effectively modelled in terms of premature/accelerated brain ageing.

When trying to disentangle disease-specific effects, the proposed DD gap metric exhibited a low correlation with BAG, supporting the relative independence between the two measures and the underlying phenomena. It should be noted that DD is an intrinsically noisy measure, relying on the date of clinical onset, which is often assigned retrospectively based on the subjective recollection of symptoms. Nevertheless, the model performance was above chance level, and explained considerably more variance in DD than other established measures of multiple sclerosis-related brain involvement such as total lesion and thalamic volumes. On the other hand, the MS-age model was highly accurate, nearly approaching the performance of state-of-the-art deep learning models of healthy brain ageing^14,26–28^, but the corresponding gap measure was highly correlated with BAG, suggesting a greater degree of residual dependence from healthy brain ageing patterns.

The interpretability analyses showed that all measures were influenced by regional brain volumes, with the lowest effect sizes observed for the MS-age gap metric, revealing the lower capacity of the corresponding model to capture inter-subject variability. In line with what has been previously observed for healthy brain ageing models,^29,30^ age and DD predictions were influenced by spatially distributed, rather than localized, variations in brain volume. Interestingly, the presence of lesions did not directly influence BAG values, while MS-age and, most prominently, DD predictions on unfilled scans were systematically higher than those obtained on lesion-filled counterparts, suggesting that multiple sclerosis-specific models can effectively measure disease-related phenomena that are not captured by the classical brain-age paradigm.

This idea was further supported by the association with physical disability, with the DD gap metric explaining additional variance in the EDSS score compared to BAG alone. Longitudinal trajectories estimated on the early multiple sclerosis cohort substantiated the biological interpretation of the investigated metrics. EDSS, as a measure of physical disability, and BAG, expressing the deviation from healthy brain ageing, tend to increase over time on average as a reflection of disease progression. Conversely, multiple sclerosis-specific metrics express deviation from the average PwMS and therefore do not exhibit significant group-level change over time.

It should be noted that early multiple sclerosis represents the ideal setting to analytically separate ageing and disease-specific effects, as the relative contribution of normal ageing to brain atrophy is low and the amount of disease-related variance in brain structure that is not explained by ageing is higher.^8^ Indeed, a ceiling effect is observable for brain structural damage, with the trajectories of brain volume change in PwMS and HS tending to align in the elderly.^31^

From the clinical perspective, the longitudinal association between BAG and physical disability has been previously demonstrated.^7^ We showed that disability worsening is also paralleled by the increase in DD gap, reflecting accelerated progression of multiple sclerosis-specific brain damage compared to reference trajectories and adding to the BAG metric in explaining EDSS change over time.

Taken together, our results show that complementing the brain-age paradigm with models explicitly designed to capture disease-specific effects allows to comprehensively measure both ageing-like and non ageing-like aspects of brain pathology, providing a more accurate explanation of brain damage and related disability in PwMS. The DD gap metric, in particular, is sensitive to imaging patterns (and underlying biological processes) that are relatively independent from healthy brain ageing and therefore adds to classical brain-age models as a disease-specific biomarker. The model of multiple sclerosis-specific ageing, on the other hand, is not as specific, probably because of its sensitivity to physiological, non disease-related variability across subjects, and does not seem to add to the classical brain-age paradigm.

Our study is not without limitations. First, the DD model is prone to biases. As mentioned, DD is an intrinsically noisy measure, with the length of the preclinical phase being influenced by several, not necessarily random, factors.^32^ For a more accurate estimation of reference disease trajectories, we need more accurate estimates of disease onset relying on objective biomarkers.^32,33^ Also, consideration is needed on the potential confounding role of disease modifying drugs. In recent years, the therapeutic landscape in multiple sclerosis has drastically changed, with the proliferation of highly effective treatment options making PwMS with a more recent diagnosis more likely to have milder disease courses. More complex models taking into account the effect of treatment as a disease course modifier will be needed to solve this possible bias. Furthermore, some caution is needed when interpreting gap values due to their DD-dependence (i.e., the underestimation of DD in PwMS with longer disease and vice versa). Similarly to healthy brain ageing models, care should be taken to apply statistical bias correction before or during downstream analyses.^34^ Second, further clinical validation with additional outcome measures, longer follow-up times and more varied, real-world, clinical populations will be needed to fully assess the potential of the DD gap metric for patient stratification in clinical practice. It is also worth noting that the realm of deep learning methods offers possible alternatives to solve the problem of unraveling brain ageing and disease-specific effects, with disentangled representation learning approaches being particularly promising in this regard and potentially representing a crucial area for future research.^35,36^

In conclusion, we demonstrated that the DD gap is a clinically meaningful measure of multiple sclerosis-specific brain damage, adding to models of healthy brain ageing. Condensing the complex information contained in routinely acquired brain MRI scans into a simple and intuitive biomarker of disease severity and progression, it may represent a powerful tool for the stratification of PwMS in both clinical and research settings.

## Appendix

Authors are members of the MAGNIMS network (Magnetic Resonance Imaging in multiple sclerosis; https://www.magnims.eu/), which is a group of European clinicians and scientists with an interest in undertaking collaborative studies using MRI methods in multiple sclerosis, independent of any other organization. The group is run by a steering committee whose members are: Frederik Barkhof (Amsterdam), Nicola de Stefano (Siena), Jaume Sastre-Garriga (Barcelona, Co-Chair), Olga Ciccarelli (London), Christian Enzinger (Graz), Massimo Filippi (Milan), Claudio Gasperini (Rome), Ludwig Kappos (Basel), Jacqueline Palace (Oxford), Hugo Vrenken (Amsterdam), Alex Rovira (Barcelona), Maria Assunta Rocca (Milan, Co-Chair), and Tarek Yousry (London).

