## Supplementary Material for "Disentangling neurodegeneration from ageing in multiple sclerosis: the brain-predicted disease duration gap"

### Supplementary Table 1. MRI acquisition protocols.

| Centre | Barcelona I | Barcelona II | Basel | Bochum | Graz | Mainz | Milan | Naples I | Naples II |
|---|---|---|---|---|---|---|---|---|---|
| Field strength | 3 Tesla | 3 Tesla | 3 Tesla | 3 Tesla | 3 Tesla | 3 Tesla | 3 Tesla | 3 Tesla | 3 Tesla |
| Vendor | Siemens | Siemens | Siemens | Philips | Siemens | Siemens | Philips | Siemens | GE |
| Model | Trio/Prisma | Trio | Skyra | Achieva | Prisma | Trio | Ingenia | Trio | Discovery |
| Years of recruitment | 2016-2022 | 2016-2019 | 2017-2020 | 2011-2019 | 2021-2022 | 2017-2019 | 2017-2020 | 2016-2022 | 2019-2022 |
| Voxel dimensions (mm) | 1x1x1 | 0.94x0.94x0.94 | 1x1x1 | 1x1x1 | 1x1x1 | 1x1x1 | 1x1x1 | 0.8x0.8x0.8 | 1x1x1 |
| TR (ms) | 2300 | 1800 | 2300 | 10 | 1900 | 1900 | 7 | 3000 | 7 |
| TE (ms) | 3 | 3 | 2 | 4.6 | 2.7 | 2.5 | 3.2 | 2.4 | 3 |
| TI (ms) | 900 | 800 | 900 | - | 900 | 900 | 1000 | 1000 | 650 |
| FA (°) | 9 | 9 | 9 | 8 | 9 | 9 | 8 | 9 | 9 |
| Slices, Orientation | 176, sagittal | 240, sagittal | 192, sagittal | 180, sagittal | 176, sagittal | 192, sagittal | 204, sagittal | 224, sagittal | 206, sagittal |

| Centre | Oslo | | | | Oxford | Prague | Rome | Siena | Verona | London I | | London II |
|---|---|---|---|---|---|---|---|---|---|---|---|---|
| Field strength | 3 Tesla | 3 Tesla | 3 Tesla | 1.5 Tesla | 3 Tesla | 3 Tesla | 1.5 Tesla | 3 Tesla | 3 Tesla | 1.5 Tesla | 3 Tesla | 3 Tesla |
| Vendor | GE | GE | GE | Siemens | Siemens | Siemens | Siemens | Philips | Philips | GE | Philips | Philips |
| Model | Signa | Discovery | Signa | Avanto | Prisma | Skyra | Avanto | Achieva | Achieva | Signa | Achieva | Achieva |
| Years of recruitment | 2012-2014 | 2015-2019 | 2019-2022 | 2012-2017 | 2018-2019 | 2012-2022 | 2018-2021 | 2017-2021 | 2015-2017 | 1999-2008 | 2014-2015 | 2014-2023 |
| Voxel dimensions (mm) | 1.2x0.5x0.5 | 1x1x1 | 0.8x0.8x0.8 | 1.2x1.25x1.25 | 1x1x1 | 1x1x1 | 1x1x1 | 1x1x1 | 1x1x1 | 1.5x1.5x0.9 | 1x1x1 | 1x1x1 |
| TR (ms) | 10.2 | 8.2 | 2356 | 2400 | 2040 | 2300 | 2200 | 10 | 8.2 | 10.9 | 6.8 | 7 |
| TE (ms) | 4.2 | 3.2 | 3 | 3.6 | 4.7 | 3 | 3.4 | 4 | 3.8 | 4.2 | 3.0 | 3.2 |
| TI (ms) | 450 | 450 | 950 | 1000 | 900 | 900 | 950 | - | - | 450 | - | - |
| FA (°) | 13 | 12 | 8 | 8 | 8 | 9 | 8 | 8 | 8 | 20 | 8 | 8 |
| Slices, Orientation | 124, axial | 186, sagittal | 240, sagittal | 160, sagittal | 192, sagittal | 176, sagittal | 176, axial | 256, sagittal | 180, sagittal | 124, coronal | 180, sagittal | 176, sagittal |

mm = millimetre; ms = milliseconds; TE = echo time; TR = repetition time; FA = flip angle.

**Supplementary Table 2. Models for the prediction of EDSS.** Coefficient estimates (with standard errors in parentheses) and model statistics for the linear regression analyses predicting EDSS.

| | *Dependent variable:* | | |
|---|---|---|---|
| | EDSS | | |
| | (1) | (2) | (3) |
| Brain-age gap | 0.026$^{***}$ | | 0.023$^{***}$ |
| | (0.005) | | (0.005) |
| MS-age gap | | 0.031$^{***}$ | 0.017 |
| | | (0.010) | (0.011) |
| Age | 13.786$^{***}$ | 12.898$^{***}$ | 14.483$^{***}$ |
| | (1.656) | (1.687) | (1.709) |
| Age$^2$ | 2.093 | 2.226 | 2.279$^{*}$ |
| | (1.337) | (1.354) | (1.340) |
| DD | 0.023$^{***}$ | 0.031$^{***}$ | 0.023$^{***}$ |
| | (0.006) | (0.006) | (0.006) |
| Sex | -0.094 | -0.046 | -0.100 |
| | (0.096) | (0.096) | (0.096) |
| Constant | 1.900$^{***}$ | 1.987$^{***}$ | 1.912$^{***}$ |
| | (0.091) | (0.090) | (0.091) |
| Observations | 867 | 867 | 867 |
| $R^2$ | 0.187 | 0.171 | 0.189 |
| Adjusted $R^2$ | 0.182 | 0.166 | 0.184 |
| Residual Std. Error | 1.318 (df = 861) | 1.331 (df = 861) | 1.317 (df = 860) |
| F Statistic | 39.531$^{***}$ (df = 5; 861) | 35.577$^{***}$ (df = 5; 861) | 33.445$^{***}$ (df = 6; 860) |

*Note:* MS = multiple sclerosis; DD = disease duration; df = degrees of freedom. *p<0.1; **p<0.05; ***p<0.01

**Supplementary Table 3. Growth models of EDSS and age and disease duration gaps in the early multiple sclerosis cohort.** EDSS, brain-age gap, disease duration gap, and MS-age gap are the dependent variables of multilevel linear models with timepoints nested within subjects and random intercept and slope of follow-up time per subject, including also the fixed effects of age, age$^2$ (to account for the non-linear effect of age), and sex. When modelling the disease duration gap, the fixed effect of disease duration was also included in the model to correct for disease duration-related bias (i.e., the underestimation of disease duration in long-standing pwMS and vice versa). Shown are the coefficient estimates (with standard errors in parentheses) and model statistics.

| | *Dependent variable:* | | | |
|---|---|---|---|---|
| | **EDSS** | **BAG** | **DD gap** | **MS-age gap** |
| | **(1)** | **(2)** | **(3)** | **(4)** |
| Follow-up time | 0.058*** | 0.472*** | 0.057 | 0.016 |
| | (0.016) | (0.089) | (0.046) | (0.052) |
| Age | 6.614*** | -33.343** | 26.833*** | -39.314*** |
| | (1.764) | (13.972) | (4.650) | (8.781) |
| Age$^2$ | 2.736* | -32.054*** | -7.353** | -18.157*** |
| | (1.397) | (9.026) | (3.177) | (5.371) |
| DD | | | -0.825*** | |
| | | | (0.042) | |
| Sex | 0.086 | 3.544*** | 0.540* | 1.163* |
| | (0.127) | (1.002) | (0.298) | (0.627) |
| Constant | 1.153*** | 3.651*** | 2.800*** | 0.189 |
| | (0.080) | (0.631) | (0.209) | (0.405) |
| Observations | 678 | 749 | 749 | 749 |
| Log Likelihood | -964.876 | -2,445.987 | -1,578.271 | -2,098.721 |
| Akaike Inf. Crit. | 1,949.751 | 4,911.975 | 3,178.543 | 4,217.443 |
| Bayesian Inf. Crit. | 1,994.943 | 4,958.162 | 3,229.349 | 4,263.630 |

*Note:* BAG = brain-age gap; DD = disease duration; MS = multiple sclerosis. *p<0.1; **p<0.05; ***p<0.01

**Supplementary Table 4. Models for the prediction of annualised EDSS change.** Coefficient estimates (with standard errors in parentheses) and model statistics for the linear regression analyses predicting annualised EDSS.

| | *Dependent variable:* | | |
|---|---|---|---|
| | **EDSS annualised change** | | |
| | **(1)** | **(2)** | **(3)** |
| Brain-age gap annualised change | 0.177*** | | 0.193*** |
| | (0.023) | | (0.026) |
| MS-age gap annualised change | | 0.162 | -0.152 |
| | | (0.104) | (0.100) |
| Constant | -0.027** | 0.054*** | -0.032** |
| | (0.012) | (0.006) | (0.013) |
| Observations | 195 | 195 | 195 |
| $R^2$ | 0.228 | 0.012 | 0.238 |
| Adjusted $R^2$ | 0.224 | 0.007 | 0.230 |
| Residual Std. Error | 0.069 (df = 193) | 0.078 (df = 193) | 0.069 (df = 192) |
| F Statistic | 57.158*** (df = 1; 193) | 2.430 (df = 1; 193) | 29.917*** (df = 2; 192) |

*Note:* MS = multiple sclerosis; df = degrees of freedom. *p<0.1; **p<0.05; ***p<0.01

**Supplementary Figure 1. Conceptual framework for the MS-age modelling strategy.** Chronological age is modelled as a function of brain MRI scans in PwMS, to estimate a reference trajectory of multiple sclerosis-specific brain ageing (MS-age). The error associated with the model predictions (the brain-predicted MS-age gap), quantifies the extent to which a patient deviates from typical multiple sclerosis-specific brain ageing.

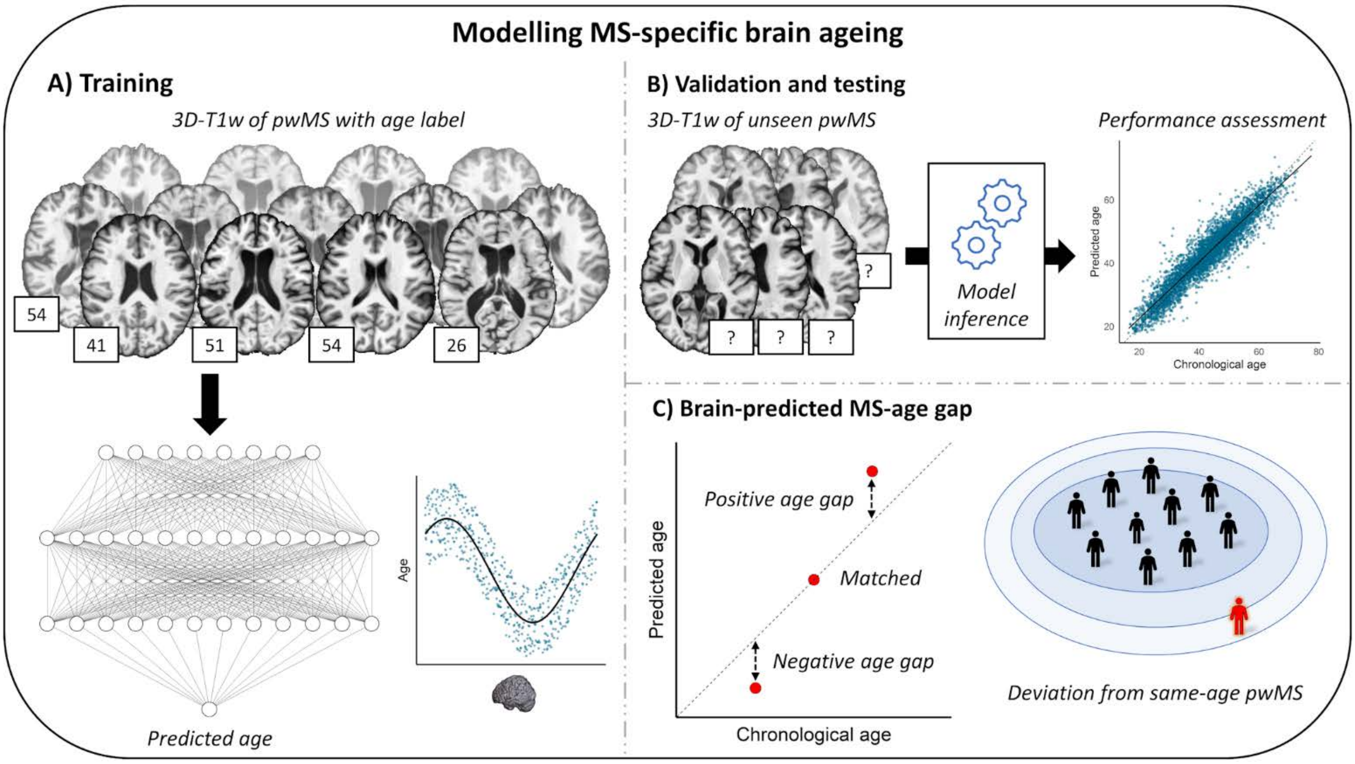

**Supplementary Figure 2. Modelling multiple sclerosis-specific brain ageing.** In (A), scatterplot showing the relationship between chronological age in the test set (N = 878) and the values predicted by the model of multiple sclerosis-specific ageing. In (B), scatterplot showing the relationship between the MS-age gap and the brain-age gap (obtained with the DeepBrainNet model) in the test set; marginal density plots are also shown, portraying the distribution of the two variables. Linear fit lines are shown as solid lines (with corresponding 95% confidence intervals in grey), while dashed lines represent the line of identity (A), and horizontal and vertical zero reference lines (B), respectively.

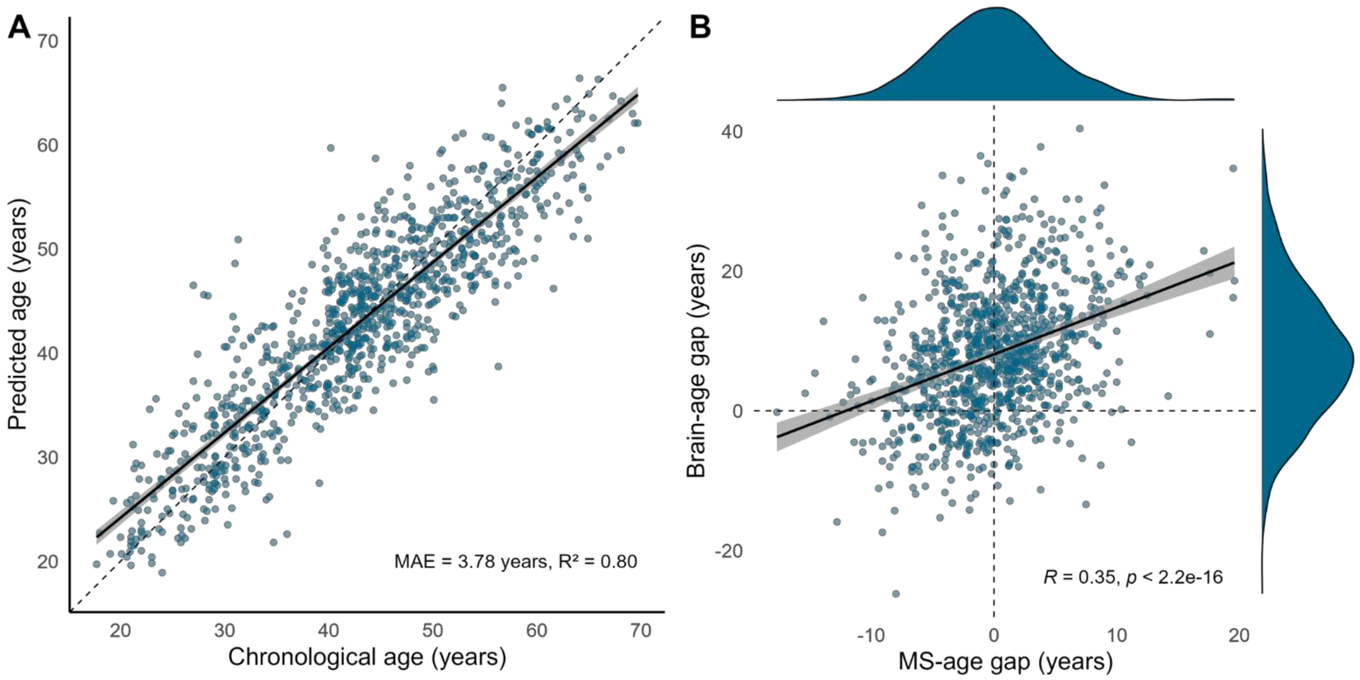

**Supplementary Figure 3. Guided backpropagation analysis to interrogate brain regions influencing the model for the prediction of MS-age.** Lightbox view of selected slices from the quasi-raw T1w volumes (on the left) and corresponding guided backpropagation-derived saliency maps (on the right) of the same subjects presented in Figure 3. For saliency maps, both positive (positively correlated with the output, in *red*) and negative (negatively correlated with the outcome, in *blue*) magnitudes are shown. In both cases, the model focuses mostly on regions that appear to be related to (the widening of) the cerebrospinal fluid spaces.

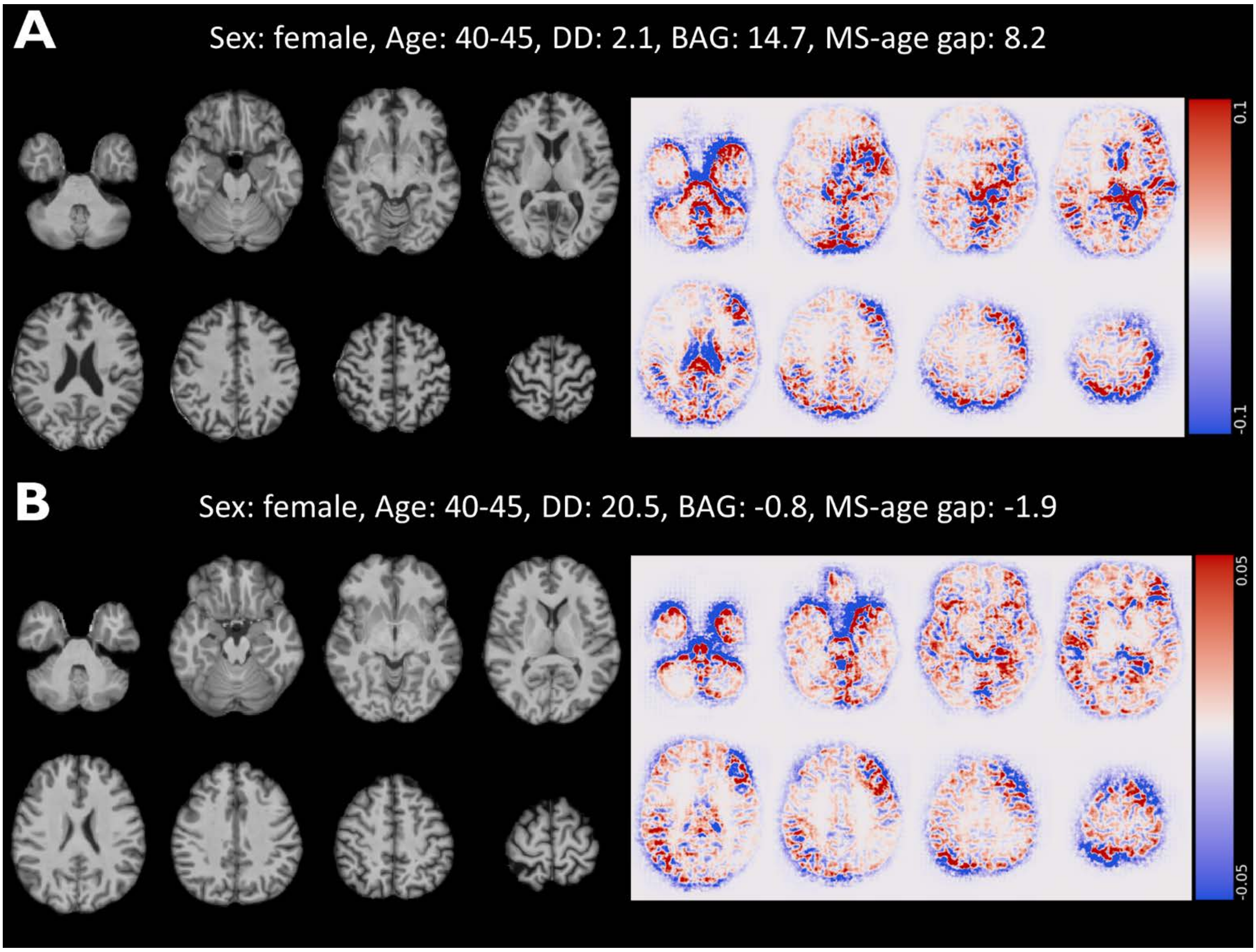

**Supplementary Figure 4. Correlations between MS-age gap and regional brain and lesion volumes.** Plots showing the correlations between MS-age gap values and cortical (A) and subcortical/lesion (B) volumes. Shown are the Pearson correlation coefficients resulting from partial correlation analyses correcting for age, age$^2$, disease duration, sex, and estimated total intracranial volume.

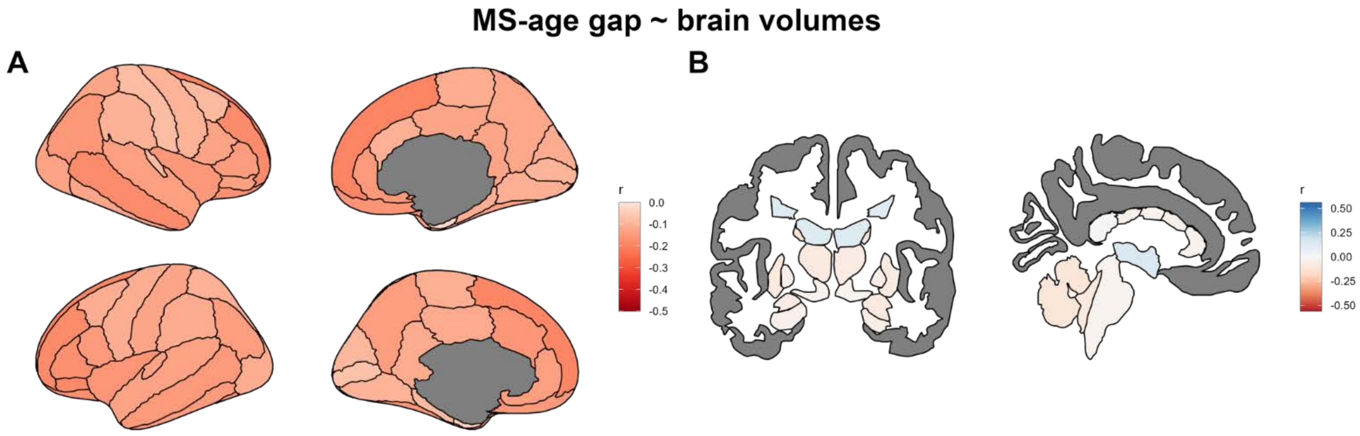

**Figure 5. Impact of MS lesions on age predictions.** Bland-Altman plot of brain-predicted MS-age (A) and age (B) from unfilled and filled T1w scans. The plots show the mean value from the 2 measures for each participant (x-axis) and the difference between the 2 measures (y-axis). The mean difference lines are solid, and the corresponding limits of agreement (±1.96 * standard deviation of difference) are dashed lines.

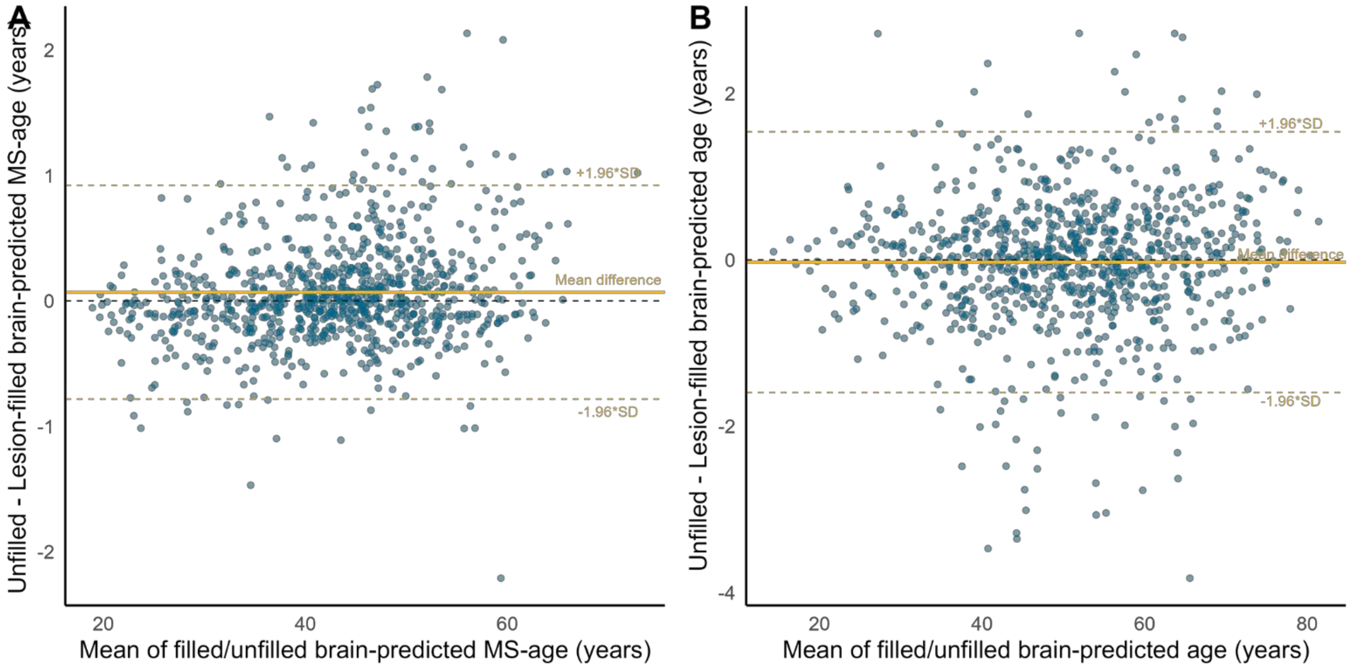

**Supplementary Figure 6. Relationships between brain-age and MS-age gaps and physical disability.** Scatterplots showing the marginal effects on EDSS of the brain-age (A) and MS-age (B) gap metrics. Regression models were corrected for the effects of age, $age^2$, disease duration, and sex. Linear fit lines are shown as solid lines (with corresponding 95% confidence intervals in grey).

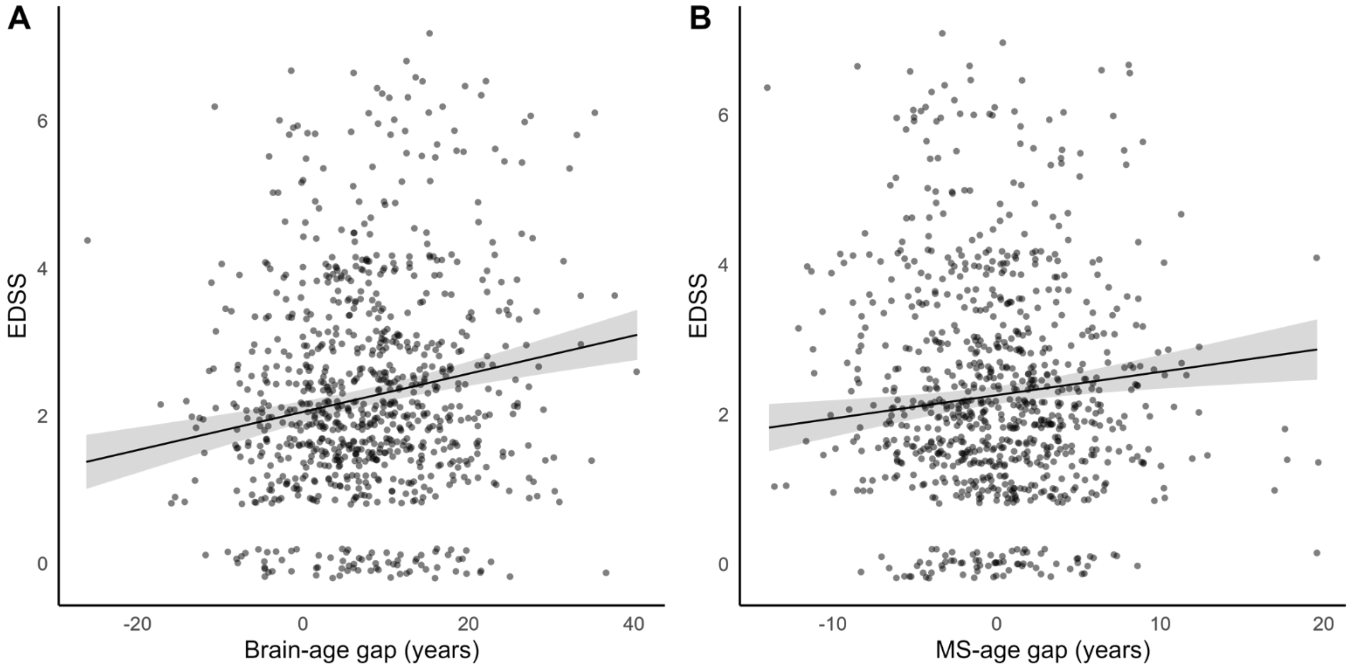

**Supplementary Figure 7. Growth models of EDSS and age and disease duration gaps in the early multiple sclerosis cohort.** Scatterplots showing the marginal effects of follow-up time on EDSS (A), brain-age gap (B), disease duration gap (C), and MS-age gap (D). Both EDSS and brain-age gap significantly increased over time, while disease duration and MS-age gaps only exhibited a slight, non-significant, upward trend. In (C), the apparent descending trend corresponding to raw data points is to be noted, mainly reflecting the bias in the disease duration prediction model (i.e., the underestimation of disease duration in long-standing pwMS and vice versa). Linear fit lines are shown as solid lines (with corresponding 95% confidence intervals in grey).

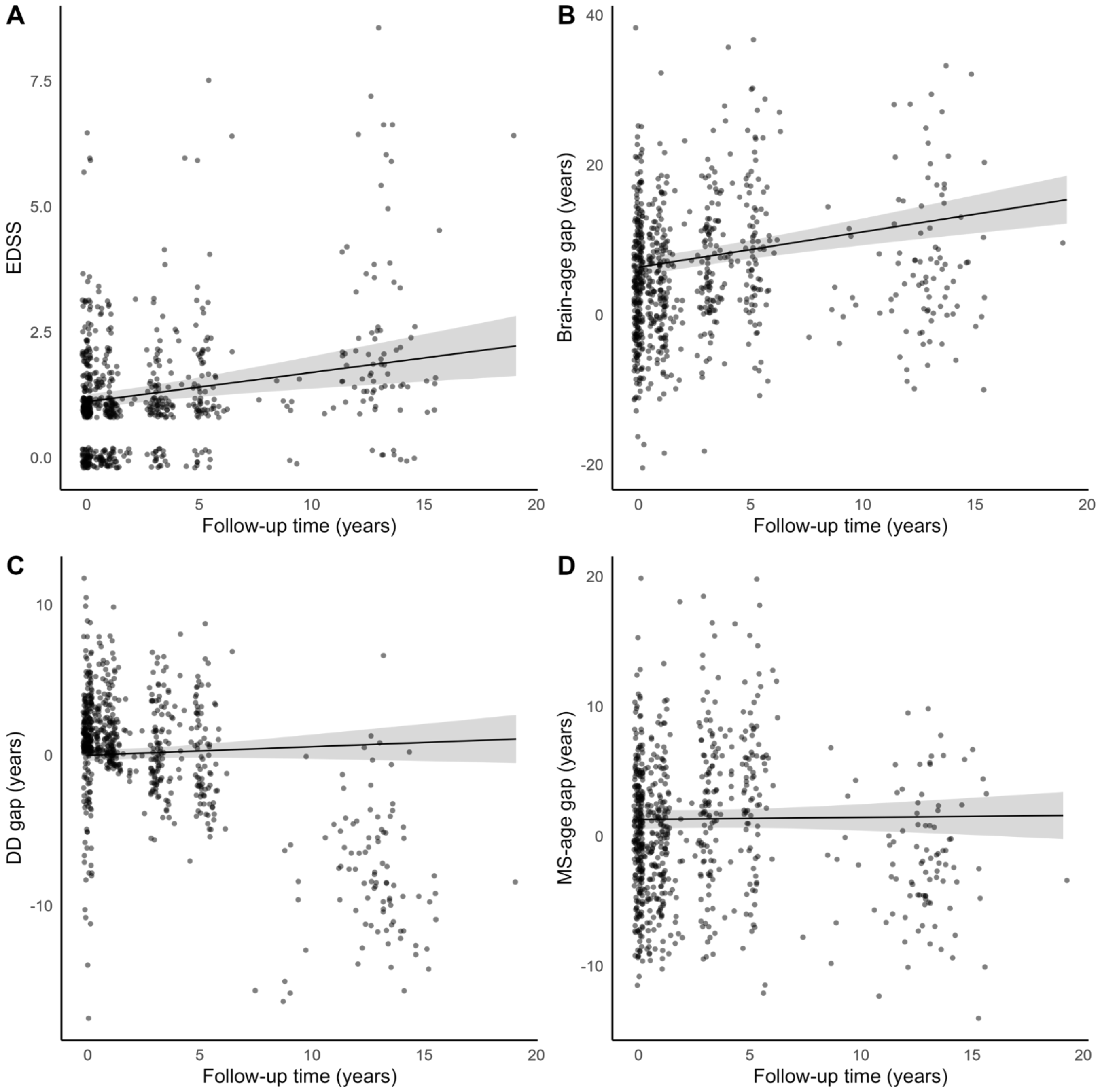

**Supplementary Figure 8. Relationships between longitudinal changes of brain-age and MS-age gaps and physical disability.** Scatterplots showing the relationship between annualised changes of EDSS and brain-age (A) and MS-age (B) gaps. Linear fit lines are shown as solid lines (with corresponding 95% confidence intervals in grey).

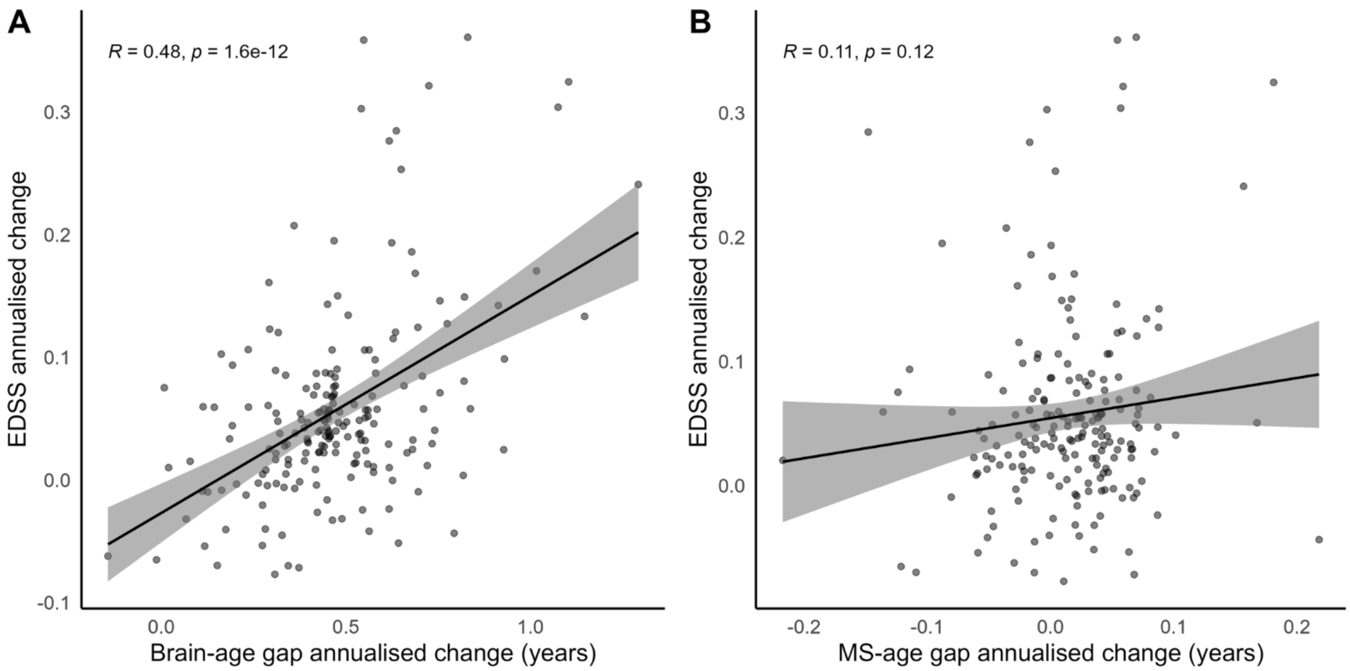
